# The prevalence of scabies, pyoderma and other communicable dermatoses in the Bijagos Archipelago, Guinea-Bissau

**DOI:** 10.1101/19000257

**Authors:** Michael Marks, Thomas Sammut, Marito Gomes Cabral, Eunice Teixeira da Silva, Adriana Goncalves, Amabelia Rodrigues, Cristóvão Manjuba, Jose Nakutum, Janete Ca, Umberto D’Alessandro, Jane Achan, James Logan, Robin Bailey, David Mabey, Anna Last, Stephen L. Walker

## Abstract

**Introduction:** Skin diseases represent a significant public health problem in most low and middle income settings. Nevertheless, there is a relative paucity of high-quality epidemiological data on the prevalence of these conditions.

**Materials/methods:** We conducted two cross-sectional population-based skin-surveys of children (6 months to 9 years old) in the Bijagós Archipelago of Guinea-Bissau during the dry season (February-March 2018) and the wet season (June-July 2018). Following a period of training, a nurse performed a standardised examination for communicable dermatoses for each participant. We calculated the prevalence of each skin condition and investigated demographic associations.

**Results:** 1062 children were enrolled in the dry season survey of whom 318 (29.9%) had at least one skin diseases. The most common diagnosis was tinea capitis (154/1062, 14.5% - 95% CI 12.5-16.8%) followed by tinea corporis (84/1062, 7.9% - 95% CI 6.4-9.7%), pyoderma (82/1062, 7.7% - 95% CI 6.2-9.5%) and scabies (56/1062. 5.2% - 95%CI 4.0 – 6.8%). 320 children were enrolled in the wet season survey of whom 121 (37.8%) had at least one skin problem. Tinea capitis remained the most common diagnosis (79/320, 24.7% - 95% CI 20.1 – 29.9%), followed by pyoderma (38/320, 11.9% - 95% CI 8.6-16.1%), tinea corporis (23/320, 7.2% - 95% 4.7 – 10.7%) and scabies (6/320, 1.9% - 95% CI 0.8-4.2%).

**Conclusions:** Our study, which utilised robust population-based cluster random sampling methodology, demonstrates the substantial disease burden caused by common communicable dermatoses in this setting. Given these findings, there is a need to consider common dermatoses as part of Universal Health Coverage to deliver ‘skin-health for all’.

**Author Summary:** Skin conditions are very common in many low and middle income settings but there have been relatively few surveys of skin disease conducted using the best epidemiological approaches. We performed two cross-sectional population-based skin-surveys of children in the Bijagós Archipelago, Guinea-Bissau, using gold-standard sampling methodologies. Skin conditions were extremely common and almost 30% of children had at least one common, infectious skin condition in both surveys. Fungal skin and scalp infections were the most common conditions followed by bacterial skin infections. Our survey demonstrates that common, easily treatable skin conditions are responsible for a high burden of disease in this population. Skin-health should be considered a key component of the Universal Health Coverage agenda.

## Introduction

The prevalence of skin disease in many low and middle income countries is high and both communicable and non-communicable skin diseases represent a significant public health problem in these settings [1]. Skin diseases are amongst the leading causes of disability adjusted life years[1]. The World Health Organization (WHO) has established programmes for Neglected Tropical Diseases (NTDs) including several severe, stigmatising skin diseases such as Buruli ulcer, leprosy and leishmaniasis. However, more common dermatoses are responsible for a significant burden of disease, in particular fungal skin infections, scabies and pyoderma which are all amongst the 50 most prevalent diseases worldwide. Common skin infections and infestations, such as scabies, are associated with significant impairment of health-related quality of life.[2].

In view of the morbidity associated with infestation, and its association with pyoderma, scabies has recently been adopted by WHO as an NTD. The majority of high quality surveys for scabies have been conducted in the Pacific or Latin America[3] with fewer data available from Africa. Data suggest that regular mass drug administration (MDA) conducted for the control of onchocerciasis or lymphatic filariasis may be responsible for a reduced prevalence of scabies in Africa but this is not a consistent finding[4,5]. Furthermore large outbreaks of scabies have been reported in Africa in recent years including an outbreak in Ethiopia involving more than one million individuals[6].

Despite the high burden of morbidity associated with these diseases there is a relative paucity of high quality epidemiological data on their prevalence. The majority of studies have used epidemiological methods which may be prone to bias such as school based surveys or screening individuals who report a skin problem[2,7,8]. As skin diseases may be associated with stigma and reduced school attendance these methods may misrepresent the true burden of morbidity due to these more common skin conditions in communities.

Since 2011 we have been conducting research on NTDs and other conditions of public health concern in the Bijagos Archipelago of Guinea-Bissau [9]. In 2018 we conducted a skin-survey of children to provide population level estimates of the prevalence of common infectious dermatoses in this population.

## Methods

### Setting

The Republic of Guinea-Bissau is located in coastal West Africa and has an estimated population of 1,792,338 with a median age of 20 years. Gross Domestic Product per capita is estimated at 1,800 USD which is among the lowest income in the world. The Bijagos Archipelago is a collection of 18 main islands and other small uninhabited islands. Bubaque is the major island of the archipelago. Health care on Bubaque is provided by a single small hospital facility alongside community healthcare provided by nursing posts and community health volunteers. The Ministry of Health (MoH) has previously conducted Mass Drug Administration (MDA) campaigns as part of programmes for the elimination of NTDs in the archipelago including azithromycin MDA for trachoma (2013-2016) and ivermectin and albendazole for lymphatic filariasis (2017-2018).

Ethical approval was obtained from the Research Ethics Committee of the London School of Hygiene and Tropical Medicine and the Comite Nacional de Etica na Saude of Guinea-Bissau. Permission was given from village leaders for the surveys to take place. Written informed consent was obtained from participants parents or guardians and, where possible, verbal assent was obtained from children.

### Diagnosis and training

We conducted a one-week intensive interactive training led by two physicians with experience in the diagnosis and management of scabies, pyoderma and other common dermatoses in Africa. As part of this exercise two registered nurses were trained to undertake standardised skin examinations and diagnose common dermatoses. We utilised both classroom and supervised field training sessions on dermatology history and examination skills with an emphasis on common skin infections. Diagnostic criteria were based on algorithms that had been previously validated in both the Pacific and Africa[10,11]. Scabies was diagnosed on the presence of pruritus and typical skin lesions including papules, vesicles and pustules in a typical distribution[12]. Pyoderma was defined as any skin lesion with evidence of pus or crusts. Infected scabies was diagnosed when scabies was present with evidence of pyoderma in the same distribution. Tinea corporis was defined as patches with overlying scale. Tinea capitis was defined as areas of scalp hair loss with associated scale.

### Study design, sampling and procedures

Two cross-sectional surveys of skin diseases were performed in the Bijagos Archipelago. The primary survey was conducted in February-March 2018 during the dry season. A probability proportionate to size methodology was used to select villages across the archipelago. In each selected village all children aged 1-9 were enrolled in the study. To assess for any evidence of seasonal variation we conducted a smaller secondary survey in the most populated island of the archipelago, Bubaque, during the wet season (June-July 2018). In the second survey we randomly selected 1 in 5 households in all 17 villages and again enrolled children in to the survey. We collected demographic information for each individual enrolled in either survey and the trained nurses undertook a standardised examination for scabies, pyoderma and fungal infections. In the dry season survey we additionally identified individuals with chronic ulcers, possibly compatible with yaws, and performed a rapid diagnostic test for yaws[13]. Where a treatable dermatoses was identified we provided treatment in the community free of charge.

### Data collection and statistical analysis

Data were collected on tablets using OpenDataKit. Data were analysed using R 3.4.2 (The R Foundation for Statistical Computing). We present count and prevalence data, with 95% confidence intervals, for each condition during both the dry season and wet season surveys. Adjusted odds ratios (AOR) for each skin dermatoses were calculated using a random-effects multivariable logistic regression model adjusting for demographic variables and clustering at the village level. Age was treated as a continuous variable. In a secondary analysis we compared the prevalence of each dermatoses between the dry season and wet season surveys. For the dry-season survey we conducted the survey alongside a trachoma survey which aimed to enrol approximately 1,000 children, whilst the wet-season survey was conducted as a stand-alone survey. We calculated that a sample size of 385 children would be powered to detect a prevalence of each of the skin conditions of 10% +/- 3%.

## Results

### Dry Season Survey

In the primary dry season survey we enrolled 1062 children of whom 508 (47.8%) were male with a median age of 5 years (IQR 3-7) across all the inhabited islands of the archipelago. Overall 268 had evidence of one skin infection (25.2%) and 50 had evidence of two or more skin infections (4.7%).

The most common diagnosis during the dry season was tinea capitis (154/1062, 14.5% - 95% CI 12.5-16.8%), followed by tinea corporis (84/1062, 7.9% - 95% CI 6.4-9.7%), pyoderma (82/1062, 7.7% - 95% CI 6.2-9.5%) and scabies (56/1062. 5.2% - 95%CI 4.0 – 6.8%). We identified 3 individuals (0.3%) with chronic ulcers but all had a negative RDT for yaws. Both tinea capitis (aOR 3.03, 95% CI 2.09 – 4.48) and tinea corporis (aOR 1.51, 95% CI 0.96-2.39) were more common in males although only the association with tinea capitis was statistically significant (p <0.01). There was no association between scabies and gender (aOR 1.29, 95% CI 0.74 – 2.25) nor age (aOR 0.89, 95%CI 0.81 –1.0). Pyoderma was strongly associated with scabies (aOR 3.1, 95% CI 1.38 – 6.5). The population attributable risk fraction for pyoderma due to scabies was 7%. Risk factor data for each condition during the dry season survey is shown in **Table 1**.

**Table 1:** Association between demographic factors and common skin dermatoses during the dry season survey

|  | Tinea Capitis |  | Tinea Corporis |  | Scabies |  | Pyoderma |  |
| --- | --- | --- | --- | --- | --- | --- | --- | --- |
|  | Odds Ratio<br>(95% CI) | Adjusted<br>Odds Ratio<br>(95% CI) | Odds Ratio<br>(95% CI) | Adjusted<br>Odds Ratio<br>(95% CI) | Odds Ratio<br>(95% CI) | Adjusted<br>Odds Ratio<br>(95% CI) | Odds Ratio<br>(95% CI) | Adjusted<br>Odds Ratio<br>(95% CI) |
| **Age**<br>**(per year)** | 1.10<br>(1.03 – 1.18)* | 1.09 (1.02 –<br>1.17)* | 0.98 (0.89 –<br>1.06) | 0.97 (0.89 -<br>1.06) | 0.9 (0.81 -<br>1.0) | 0.89 (0.81 –<br>1.0) | 0.90 (0.82 –<br>0.99)* | 0.90 (0.82 –<br>0.99)* |
| **Male**<br>**gender** | 3.03 (2.09-4.48)** | 2.97 (2.05 –<br>4.38)** | 1.51 (0.96 –<br>2.39) | 1.53 (0.97 –<br>2.43) | 1.24 (0.75 –<br>2.15) | 1.29 (0.74 –<br>2.25) | 1.45 (0.91 –<br>2.3) | 1.48 (0.93 –<br>2.38) |
| **Presence of**<br>**Scabies** | N/A | N/A | N/A | N/A | N/A | N/A | 3.31 (1.47 –<br>6.89)** | 3.1 (1.38 –<br>6.5)** |

\* p < 0.05
\*\* p < 0.01

### Wet Season Survey

In the wet season survey we enrolled 320 children of whom 162 (50.6%) were male with a median age of 5 years (IQR 3-7). Overall 99 children had evidence of one skin infection (30.9%) and 22 children had evidence of two or more skin infections (6.9%).

During the wet season tinea capitis remained the most common diagnosis (79/320, 24.7% - 95% CI 20.1 – 29.9%), followed by pyoderma (38/320, 11.9% - 95% CI 8.6-16.1%), tinea corporis (23/320, 7.2% - 95% 4.7 – 10.7%) and scabies (6/320, 1.9% - 95% CI 0.8-4.2%). Male gender remained associated with the presence of tinea capitis (aOR 4, 95% CI 2.43 -6.00). No variables were significantly associated with either tinea corporis or scabies in the wet season survey. Increasing age was associated with a decreased risk of pyoderma (aOR per year of age 0.82, 95%CI 0.74 – 0.9). Scabies remained associated with pyoderma (aOR 3.0, 95% CI 0.04 – 15.04). The population attributable risk fraction for pyoderma due to scabies during the wet season survey was 3.5%. Risk factor data for each condition during the wet season survey is shown in **Table 2**. The prevalence of both tinea capitis (24.7% vs 14.5%, p <0.001) and pyoderma (11.9% vs 7.9%, p = 0.03) was significantly higher, whilst the prevalence of scabies was lower (1.9% vs 5.2%, p = 0.02) during the wet season survey compared to the dry season survey. The prevalence of each disease by age, gender and season are shown in **Table 3**.

**Table 2:** Association between demographic factors and common skin dermatoses during the wet season survey

|  | Tinea Capitis |  | Tinea Corporis |  | Scabies |  | Pyoderma |  |
| --- | --- | --- | --- | --- | --- | --- | --- | --- |
|  | Odds Ratio<br>(95% CI) | Adjusted<br>Odds Ratio<br>(95% CI) | Odds Ratio<br>(95% CI) | Adjusted<br>Odds Ratio<br>(95% CI) | Odds Ratio<br>(95% CI) | Adjusted<br>Odds Ratio<br>(95% CI) | Odds Ratio<br>(95% CI) | Adjusted<br>Odds Ratio<br>(95% CI) |
| **Age**<br>**(per year)** | 0.95 (0.9 – 1.00) | 0.95 (0.89 – 1.00) | 1.07 ( 0.98 – 1.15) | 1.07 (0.98 – 1.16) | 0.91 (0.74 – 1.08) | 0.91 (0.74. – 1.08) | 0.82 (0.73 – 0.89)** | 0.82 (0.74 – 0.90)* |
| **Male**<br>**gender** | 4.00 (2.43 – 6.82)** | 4.09 (2.48 – 6.98)** | 0.60 (0.31 – 1.15) | 0.59 (0.03 – 1.13) | 1.51 (0.37 – 7.41) | 1.56 (0.38 – 7.67) | 0.98 (0.52 – 1.88) | 1.01 (0.52- 1.96) |
| **Presence of**<br>**Scabies** | N/A | N/A | N/A | N/A | N/A | N/A | 3.43 (0.48 – 16.2) | 3.00 ( 0.40 – 15.04) |

**Table 3:** Prevalence of each skin infection by age, gender and season

|  |  | Dry Season |  |  |  |  |  | Wet Season |  |  |  |
| --- | --- | --- | --- | --- | --- | --- | --- | --- | --- | --- | --- |
| Age | Gender | Total | Tinea Capitis<br>n (%) | Tinea Corporis<br>n (%) | Scabies<br>n (%) | Pyoderma<br>n (%) | Total | Tinea Capitis<br>n (%) | Tinea Corporis<br>n (%) | Scabies<br>n (%) | Pyoderma<br>n (%) |
| **0** | Female | 30 | 0 (0) | 0 (0) | 1 (3.4) | 2 (6.7) | N/A | N/A | N/A | N/A | N/A |
|  | Male | 14 | 0 (0) | 1 (7.2) | 0 (0) | 2 (14.3) | N/A | N/A | N/A | N/A | N/A |
| **1** | Female | 44 | 3 (6.9) | 6 (13.7) | 4 (9.1) | 6 (13.7) | 12 | 0 (0) | 0 (0) | 0 (0) | 2 (16.7) |
|  | Male | 38 | 5 (13.2) | 4 (10.6) | 2 (5.3) | 4 (10.6) | 14 | 1 (7.2) | 1 (7.2) | 2 (14.3) | 2 (14.3) |
| **2** | Female | 53 | 1 (1.9) | 3 (5.7) | 1 (1.9) | 1 (1.9) | 18 | 2 (11.2) | 2 (11.2) | 0 (0) | 5 (27.8) |
|  | Male | 55 | 8 (14.6) | 6 (11) | 5 (9.1) | 5 (9.1) | 15 | 5 (33.4) | 0 (0) | 0 (0) | 3 (20) |
| 3 | Female | 76 | 7 (9.3) | 5 (6.6) | 6 (7.9) | 9 (11.9) | 15 | 3 (20) | 0 (0) | 0 (0) | 2 (13.4) |
|  | Male | 54 | 12 (22.3) | 10 (18.6) | 5 (9.3) | 6 (11.2) | 19 | 5 (26.4) | 0 (0) | 0 (0) | 1 (5.3) |
| 4 | Female | 60 | 7 (11.7) | 2 (3.4) | 6 (10) | 7 (11.7) | 18 | 2 (11.2) | 3 (16.7) | 1 (5.6) | 5 (27.8) |
|  | Male | 55 | 14 (25.5) | 4 (7.3) | 4 (7.3) | 6 (11) | 14 | 6 (42.9) | 2 (14.3) | 0 (0) | 4 (28.6) |
| 5 | Female | 64 | 4 (6.3) | 8 (12.5) | 2 (3.2) | 2 (3.2) | 24 | 3 (12.5) | 1 (4.2) | 1 (4.2) | 2 (8.4) |
|  | Male | 48 | 12 (25) | 2 (4.2) | 2 (4.2) | 8 (16.7) | 27 | 7 (26) | 2 (7.5) | 1 (3.8) | 1 (3.8) |
| 6 | Female | 70 | 10 (14.3) | 4 (5.8) | 3 (4.3) | 3 (4.3) | 16 | 3 (18.8) | 2 (12.5) | 0 (0) | 2 (12.5) |
|  | Male | 64 | 14 (21.9) | 5 (7.9) | 4 (6.3) | 4 (6.3) | 19 | 7 (36.9) | 2 (10.6) | 0 (0) | 2 (10.6) |
| 7 | Female | 53 | 7 (13.3) | 0 (0) | 3 (5.7) | 4 (7.6) | 20 | 3 (15) | 2 (10) | 0 (0) | 0 (0) |
|  | Male | 57 | 17 (29.9) | 6 (10.6) | 3 (5.3) | 2 (3.6) | 17 | 5 (29.5) | 0 (0) | 0 (0) | 2 (11.8) |
| 8 | Female | 43 | 4 (9.4) | 0 (0) | 1 (2.4) | 0 (0) | 19 | 2 (10.6) | 1 (5.3) | 0 (0) | 1 (5.3) |
|  | Male | 63 | 16 (25.4) | 6 (9.6) | 3 (4.8) | 4 (6.4) | 26 | 16 (61.6) | 2 (7.7) | 0 (0) | 2 (7.7) |
| 9 | Female | 61 | 4 (6.6) | 8 (13.2) | 0 (0) | 2 (3.3) | 16 | 3 (18.8) | 2 (12.5) | 0 (0) | 1 (6.3) |
|  | Male | 60 | 9 (15) | 4 (6.7) | 1 (1.7) | 5 (8.4) | 11 | 6 (54.6) | 1 (9.1) | 1 (9.1) | 1 (9.1) |

## Discussion

Our study shows the substantial burden of disease caused by common communicable dermatoses amongst children of the Bijagos Archipelago of Guinea-Bissau. In both the wet and dry season survey almost 30% of children had evidence of at least one communicable dermatosis highlighting the extent to which these easily treatable diseases are ubiquitous in this setting. A particular strength of this study is the robust, population-based cluster random sampling methodology that was used. The majority of similar studies to date have used less epidemiologically robust techniques which are at considerable risk of bias, possibly resulting in inaccurate prevalence estimates. By contrast our study used a gold-standard sampling methodology used in large-scale NTD mapping studies such as the Global Trachoma Mapping Project[14].

Fungal dermatoses were the most common dermatoses, in keeping with the findings of previous studies conducted elsewhere [1]. As previously reported[15], tinea capitis was more common amongst boys. The prevalence of scabies was relatively low compared to other studies in Africa[16]. This may, in part, be due to the round of MDA with ivermectin for the elimination of lymphatic filariasis carried out by the Ministry of Health in the previous year, which may have reduced the prevalence of scabies at this time[5,17–19]. There was a substantial burden of pyoderma in these communities and some evidence that this increased during the wet season. In other settings there is evidence that pyoderma is a driver of more serious bacterial infections, glomerulonephritis and rheumatic fever[20]. Although there is no specific data from Guinea-Bissau these sequelae including invasive *Staphylococcus aureus* and rheumatic fever are common in West Africa[21–23]. It is plausible that the burden of pyoderma in our current surveys may also be a driver for these conditions in Guinea-Bissau.

Our study has some limitations. Firstly, we examined only children aged nine years old or less. It is plausible that a different range of skin conditions are more common in older children and further work is required to explore this. Secondly, our study was limited to rural areas and should not be taken to be representative of peri-urban or urban populations in other regions of Guinea-Bissau. Our definition of tinea capitis was based on patches of hair loss. We might therefore have missed some cases of tinea capitis presenting with less common manifestations, such as kerion, but this is unlikely to significantly have affected our results. Whilst diagnoses in our study were made by a trained health worker and not by a dermatologist several studies have previously shown that a high degree of diagnostic accuracy can be achieved with training [10,11,24]; we are therefore confident that our clinical data are accurate.

WHO has proposed an integrated-approach to screening for NTDs involving the skin and has developed initial healthcare worker training materials to support this strategy [25]. Initial small scale pilot studies have demonstrated clearly that these rarer, if more serious diseases, are dwarfed by the burden of disease of common skin conditions[7,8]. Whilst this might be considered a negative, from the perspective of identifying cases of NTDs, we argue that this is in fact a positive. First, common dermatoses represent differential diagnoses of rarer NTDs of the skin and training health care workers in their recognition is vital to ensure robust diagnoses. Secondly, by identifying more common skin problems integrated approaches can contribute to universal health coverage. Thirdly, focusing on the broader conditions affecting communities is likely to positively influence community acceptability of such screening strategies. Finally, given the burden caused by these common dermatoses, they deserve treatment in their own right as part of Universal Health Coverage to deliver ‘skin-health for all’[26].

## Data Availability

Data is available in the supplementary data file.

## Acknowledgements

We extend our thanks to colleagues in the Ministério de Saúde Pública in Bissau and to the communities of the Bijagós Archipelago, our field research team and study participants.

## Supporting Information Legend

S1 Checklist – Strobe Checklist

S2 Data – Supporting Dataset

